# Beyond Boundaries: A Scoping Review of Adolescent Physical Activity in Asia and South Asia

**DOI:** 10.1101/2025.04.03.25325221

**Authors:** Saleema Gulzar, Jawaria Mukhtar Ahmed, Sharmeen Akram, Alishah Aziz

## Abstract

**Background:** Physical activity (PA) plays a crucial role in adolescent health and development. The World Health Organization recommends that adolescents engage in at least 60 minutes of moderate to vigorous PA daily, with schools being a key setting for fostering active behaviors. However, PA levels among adolescents in Low- and Middle-Income Countries (LMICs), including Pakistan, remain suboptimal, with multiple socio-cultural, environmental, and policy-related barriers influencing participation.

**Objective:** This scoping review explores the levels of adolescent PA in LMICs, particularly in Asian and South Asian middle schools, and identifies key factors affecting engagement in school-based PA.

**Methods:** Following the Arksey and O’Malley framework, we conducted a comprehensive literature search across PubMed, ERIC, CINAHL, and Embase, supplemented with grey literature sources. Studies focusing on PA levels among adolescents (ages 5 to 24 ) in LMIC school settings were included. Data were synthesized into thematic categories.

**Results:** The review highlights declining PA levels as adolescents transition from childhood, with gender disparities showing lower participation among girls. Key barriers include outdated PE curricula, lack of infrastructure, insufficiently trained PE teachers, family and cultural constraints, safety concerns, and the absence of PA-friendly policies. Gender norms, dress codes, and limited parental support further restrict engagement, particularly for girls.

Despite the well-established physical, cognitive, and socio-emotional benefits of PA, school systems in LMICs often prioritize academic performance over physical fitness, contributing to sedentary lifestyles.

**Conclusion:** Addressing the declining PA trends among adolescents requires multi-sectoral interventions, including revising PE curricula, improving infrastructure, promoting gender-inclusive PA programs, and fostering parental and community support. Evidence-based policies tailored to LMIC contexts can play a critical role in enhancing adolescent engagement in PA and mitigating long-term health risks associated with inactivity.

## Introduction

The World Health Organization has recommended that young adolescents should be engaged in 60 minutes of moderate to vigorous physical activity (PA) to maintain health (1). At least 50% of the recommended time for PA should be invested within a school setting (2) where students spend much of their time (3). Physical activity among low-middle-income countries (LMIC) adolescent students has received less attention than in the Global North. As these countries transition from low to middle-income status, there is pressure on children and families to achieve high educational outcomes at the expense of other priorities, such as preventive health behaviors and physical fitness. At the same time, low physical activity in young adolescents coincides with them becoming more engaged with their studies and scholarly outcomes (4).

Physical activity levels during adolescence have been studied extensively in non-Asian countries such as Australia, Canada, the UK, and the United States of America. A temporal trend has been noted for the sedentary lives of young adolescents aged 12-15 years. One study indicated that more than 50% of adolescents were not sufficiently active; in addition, the trend of leisure time sedentary behaviors remained very high (>40%) among adolescents (5). Overall, studies from high, middle, and low-income countries find less-than-optimal levels of physical activity for young adolescents. However, less is known about the factors that might influence physical activity levels among young adolescent students in LMIC countries such as Pakistan.

Given the pressure to perform academically, engaging in any physical activity may be considered an act of leisure (6). The pervading belief that physical activity is not useful to young adolescent development and is less important than academic studies exacerbates the problem (6, 7). A study conducted in Pakistan revealed that most physical education (PE) teachers had a positive attitude towards physical education in the school curriculum. The participants reported that physical activity is compromised because of a) insufficient budgets, b) space restrictions, c) no facilities, and d) no trained staff, indicating little system/institutional support (8).

Despite numerous well-known health benefits, physical activity levels remain lower than is recommended. Based on the literature reviewed, a scoping review was designed to determine levels of physical activity among young adolescents and identify what is known about factors that are restricting levels of physical activity in LMIC schools. The methods used to conduct the review are detailed in the following section.

## Methods

The Arksey and O’Malley framework was employed to examine the levels of and barriers to improving physical activity in LMIC schools (9). Four databases were used: PubMed, ERIC, CINAHL, and Embase. A combination of keywords and concepts were used: ‘physical activity’, ‘adolescent’, ‘youth’, teenage’, ‘school children’, ‘secondary level students’, ‘school-based physical activity’ and reports, internet sites, peer-reviewed journal articles, and professional associations included. An example search string used was “Physical activity“[tiab] AND (adolescent*[tiab] OR child*[tiab]) AND (“Physical education” OR “Physical Training”) NOT (sick[tiab] OR disable*[tiab] OR hospitalize*[tiab] OR Autism*[tiab] OR retard*[tiab]). The search strategy for grey literature used keywords in each website from the following: The survey of Pakistan’s health sector; demographic health survey Pakistan; and the World Health Organization. The review followed a two-level process. For Level 1, all the relevant abstracts were screened by the primary investigator.

Then, the full-text articles were screened for their eligibility by all authors in the Level 2 process. After selecting eligible articles, other sources were identified for possible inclusion. Figure 1 provides a detailed search strategy using the PRISMA flowchart.

**Figure 1.**
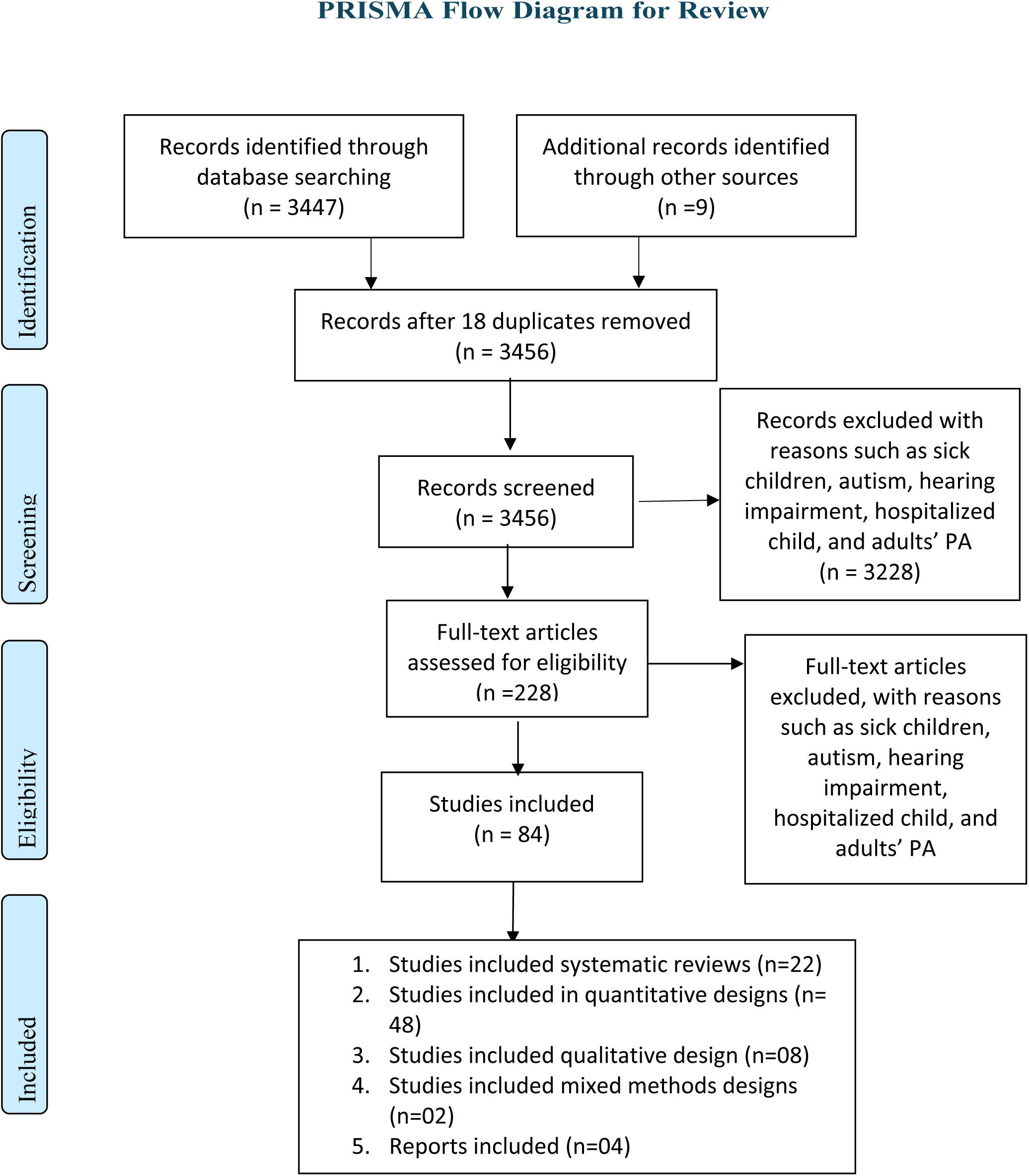
Search Strategy for Adolescent and Children Physical Activity *Search Strategy for Adolescent Physical Activity*

### Study Selection

All papers/documents were scanned by topic, and potential journal articles were retrieved. All articles were screened by the primary investigator regardless of research type and year of publication. All the cited references were imported into Endnote 20 referencing software. Relevant data tables were developed and reviewed by two other co-investigators, and a total of eighty-four were used for the final review. A comprehensive and inclusive review was conducted on all the selected manuscripts and not just those that met the standards of the appraisal tool used.

### Data Extraction and Synthesis

The duplicate articles were removed after bringing all the articles together from the databases. Articles were further sorted by titles and abstracts based on pre-defined eligibility criteria (see Figure 2). Finally, the full-text articles were identified, checked for eligibility, and then either included or excluded.

**Figure 2.**
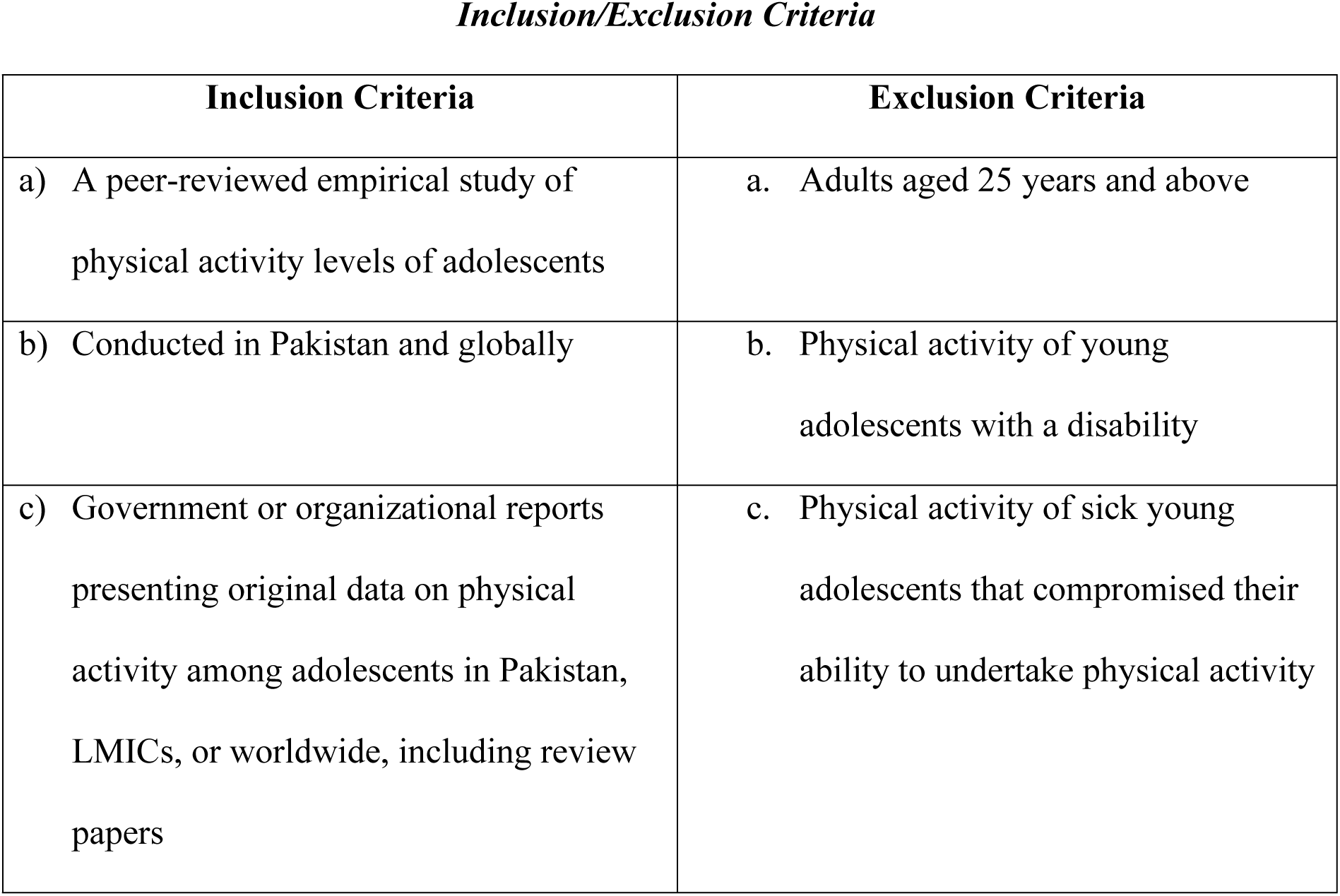
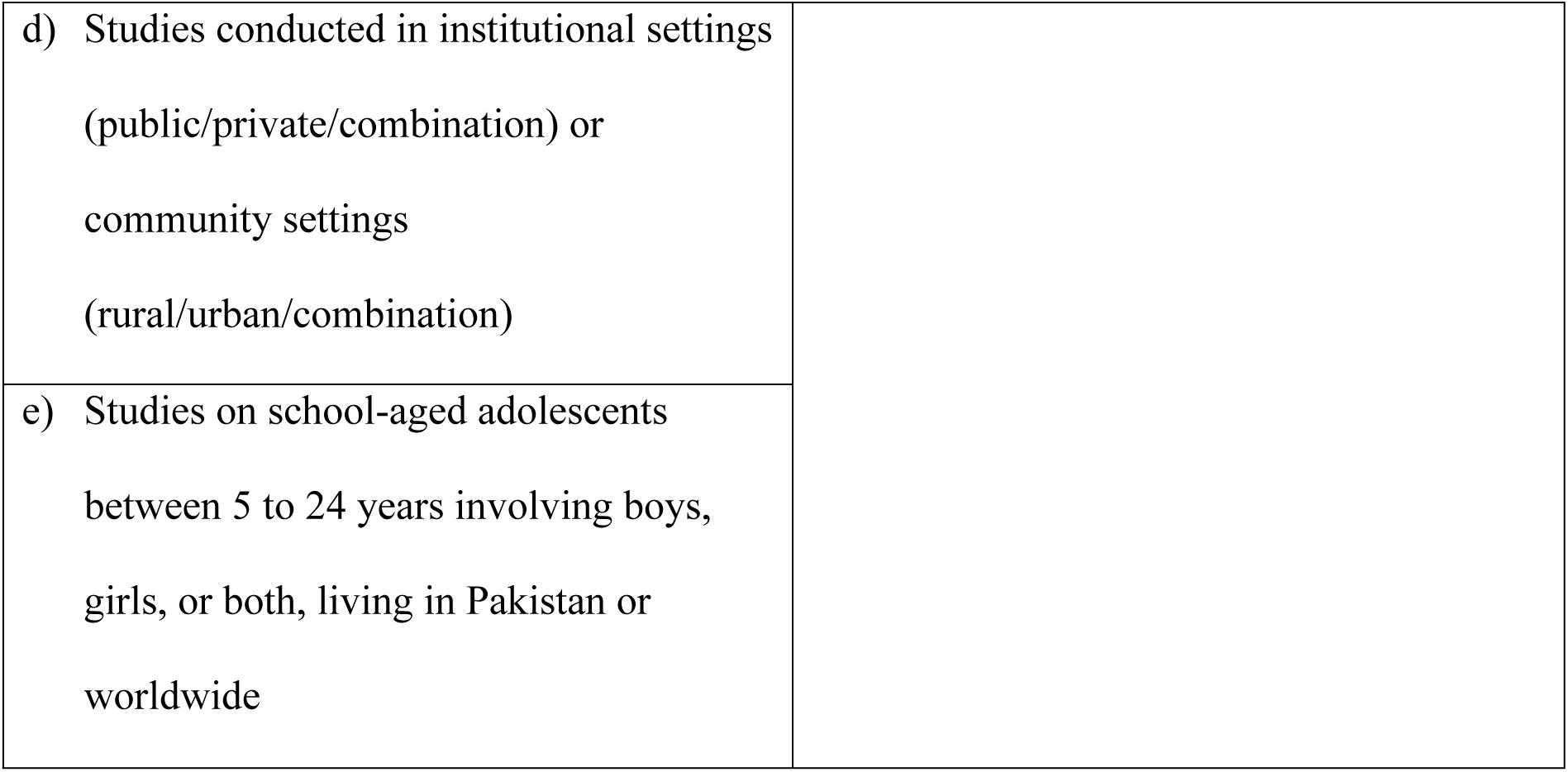
Inclusion/Exclusion Criteria

### Results and Analysis

A total of 86 peer-reviewed articles were identified, and this review is organized based on the following three major themes: 1) physical activity and adolescents, 2) benefits of physical activity, and 3) factors influencing physical activity. However, the third theme on factors affecting physical activity was further divided into several subthemes that consist of physical education curriculum, the role of physical education teachers, family support system, the role of gender, culturally appropriate dress, safety and security, and the role of infrastructure for physical activity.

#### Theme 1: Physical Activity and Adolescents

Research consistently shows a decline in physical activity levels with age as children transition into adolescence (10, 11, 12). A study in Boston, USA, highlighted a significant drop in physical activity during early to late adolescence (10). In Finland, a longitudinal study revealed that adolescents identifying as evening-oriented had lower physical activity levels (13). Another longitudinal analysis across Europe and non-European countries found declining trends in both physical activity and motor skills among youth, with increased sedentary behavior linked to higher BMI and reduced fitness (14).

A systematic review analyzing studies globally confirmed that physical activity decreases for both boys and girls as early as nine years, with a steeper decline for girls, especially during puberty (12, 15, 16, 17). Studies have shown that girls’ physical activity levels decline during the early adolescent phase, whereas, for boys, physical activity levels largely decrease in the late adolescent period (13, 18). In Pakistan, the research landscape on adolescent physical activity remains limited and inconclusive. A systematic review from January to November 2019 found low levels of physical activity among school-aged adolescents (10-19 years) but noted significant gaps in research quality, which hinder a comprehensive understanding of physical activity behavior in this population. This lack of evidence poses a critical challenge to the health and well-being of future generations in Pakistan (16).

In Pakistan, research is limited, but existing studies indicate low levels of physical activity among adolescents. For example, a study in Karachi found that 70% of secondary school students engaged in moderate activity levels, while 30% reported low activity levels, with none meeting high activity standards (19). A meta-analysis showed significant variation in reported activity levels, with a pooled prevalence of 36%, compared to the 15.5% estimated by the Pakistan Global School Health Survey (16). This review included the studies irrespective of low-quality indicators. Besides this, included studies have not adjusted key confounding variables, and therefore, generalizability could not be guaranteed.

Barriers in Pakistan include limited awareness, family support, and access to resources, along with time and financial constraints (20). The findings show a consistent decline in physical activity as children enter adolescence, with variations influenced by circadian preference, gender, and developmental stage. Internationally, physical activity drops for both boys and girls during puberty, especially for girls. In Pakistan, adolescent physical activity levels are low, with limited research leading to an incomplete understanding of the issue. Barriers like lack of awareness, resources, and family support further limit activity. Despite moderate activity in some cases, many adolescents remain insufficiently active, highlighting the need for targeted interventions to address these barriers and promote consistent physical activity.

#### Theme 2: Benefits of Physical Activity

The benefits of physical activity for adolescents span health and educational domains. Incidental activities (IA), such as household chores or walking, contribute positively to physical health and mental well-being without requiring structured time. A study in Germany among adolescents aged 12-17 found that incidental activities boosted energy and well-being, while sports improved mood but reduced calmness (17). A systematic review also linked higher physical activity levels to better health-related quality of life, while increased sedentary behavior led to poorer outcomes, especially among young adolescents (21).

Physical activity has also been shown to promote social and emotional development in adolescents. A scoping review highlighted that physical education (PE) supports social-emotional learning, improving emotional health in children aged 5-18 years (22). Further, integrating social-emotional learning competencies within PE programs fosters a positive environment that enhances students’ emotional management (23).

In Pakistan, limited interventional studies have aimed to improve adolescent physical activity. A pilot trial in Aga Khan schools involving 982 students aged 9-11 showed that interactive health education, combined with regular PE sessions, positively impacted physical activity levels and cardiometabolic health indicators like BMI and blood pressure (6). The benefits of physical activity extend across health and educational domains for adolescents. A cluster intervention trial in Karachi’s public schools highlighted improvements in blood pressure (BP) and body mass index (BMI) among 9-11-year-old girls, showing the feasibility of culturally appropriate programs in enhancing physical activity (24). However, this study’s focus on pre-teens leaves the more vulnerable adolescent girls, who globally report lower activity levels, understudied.

Educational benefits are equally significant. A systematic review involving 137 studies showed that physical activity enhances cognitive function and academic performance among children aged 5-13 years (25, 26). In Pakistan, a systematic review noted that while sleep is rarely studied, a combination of high physical activity, low sedentary behavior, and sufficient sleep yields optimal outcomes in health, psychological well-being, and education (27). In Melbourne, a study using ecological momentary assessment found that recreational moderate-to-vigorous physical activity (MVPA) boosts positive mood and energy in adolescents, though increased activity may raise tension (28).

Further, a Chinese study demonstrated that adequate physical activity, coupled with reduced sedentary behavior, positively impacts health-related quality of life (29). Recent reviews and meta-analyses confirm that structured physical education (PE) and behavior change interventions significantly enhance academic outcomes by positively influencing cognitive, affective, and psychomotor learning (30, 31). These findings underscore the dual health and educational benefits of physical activity, calling for integrated interventions targeting adolescents. This theme highlights the multifaceted benefits of physical activity for adolescents, encompassing health, educational, and social-emotional domains. Incidental activities, such as household chores and walking, contribute positively to physical and mental well-being. Structured physical activity, including sports and physical education, has been shown to improve mood, reduce stress, and enhance cognitive function. Moreover, physical activity is linked to better health-related quality of life, social-emotional development, and academic performance. Interventions aimed at promoting physical activity among adolescents, such as interactive health education and behavior change programs, have demonstrated positive outcomes in terms of physical health, mental well-being, and educational achievement.

#### Theme 3: Factors Influencing Adolescent Physical Activity

Various factors influence adolescent physical activity levels, both globally and in Pakistan. Key factors include physical education programs, structured sports, the role of teachers, family support, gender dynamics, safety concerns, and infrastructure. An analysis of these influences is presented below.

##### a. Physical Education Curriculum

Research on school-based physical education (PE) in Pakistan and internationally highlights several issues. In Pakistan, 84% of respondents view the PE curriculum as outdated, and 90% report minimal career opportunities in the field (32). Comparisons with British PE practices suggest that Pakistan could benefit from a more comprehensive approach that includes physical, emotional, and moral development, with improved teacher training and increased PE lessons (33).

Guidelines suggest 2-3 hours of PE per week to enhance physical activity among students (34, 35). Research shows that varying PE guidelines contribute to differences in adolescents’ physical activity levels across countries (36). Despite existing gaps, there is a growing interest in improving PE systems in Pakistan (8). Studies highlight the impact of physical education (PE) time allocation on adolescent physical activity (PA). In Latin America, PE days led to a 24% increase in recommended PA levels compared to non-PE days (37).

Research on African American high school girls showed that a PE intervention improved in-class PA and reduced screen time but did not increase overall daily energy expenditure. This suggests that combining PE with additional strategies, such as life skills training, enhances PA behavior (38). Increased PE time has been linked to improved student learning outcomes without harming academic performance (31). In Pakistan, insufficient awareness, resources, and opportunities contribute to a low standard of PE. Recommendations include enhancing resources, updating curricula, and increasing funding and faculty training (39). European studies reveal diverse physical literacy practices, influenced by various challenges, but show promise for future growth (40). A review of 48 studies found mixed results on the relationship between PE and academic achievement, with no evidence that increased PE time negatively impacts learning (41).

The analysis of existing literature reveals several challenges and opportunities related to physical education (PE) curricula in Pakistan. The outdated curriculum and limited career prospects in the field highlight the need for significant reforms. Comparisons with international practices suggest that Pakistan could benefit from a more comprehensive approach that includes physical, emotional, and moral development. Increasing the allocation of time for PE lessons is crucial to enhancing physical activity levels among students. While studies have shown the positive effects of PE on physical activity and academic outcomes, there is a need for further research to address the specific challenges and opportunities in the Pakistani context.

##### b. Role of Physical Education Teachers

The research underscores the pivotal role of physical education (PE) teachers in influencing students’ physical activity (PA) motivation. Studies since the early 1990s have consistently shown a positive link between effective PE teaching methods and increased student motivation for PA (42). In Pakistan, a study revealed that 10% of respondents identified a lack of training opportunities for PE teachers as a significant issue (8). Another study highlighted that controlling teaching styles, including pressuring techniques to enforce teacher-prescribed behaviors, can demotivate students (43). Authoritative teaching methods are counterproductive, whereas supportive and positive interactions enhance students’ engagement in PA (43). Research also highlights that innovative teaching strategies, such as multimedia and group discussions, are valued but often underutilized due to resource constraints (44).

The global emphasis on school-based PE over the past 20 years has highlighted the importance of providing professional development opportunities for PE teachers, as seen in the UK (45). Such professional development is linked to increased student interest in PA (46). Additionally, transformational leadership by PE teachers significantly boosts student satisfaction and enjoyment in PE classes, with task-oriented motivational styles mediating these effects (47). Recent studies on professional development for physical education (PE) teachers reveal mixed results. A study comparing two continuous professional development (CPD) methods—workshop only and workshop plus lesson study—found both approaches significantly enhanced teachers’ autonomy support and slightly improved instructional structure. However, the addition of the lesson study component did not offer additional benefits over the traditional workshop in terms of student motivation and perceived motivational climate (48).

An umbrella review of 17 pedagogical models, involving over 22,000 students and 1,000 teachers, confirmed that models such as Sport Education, Games-Centered Approach, and Cooperative Learning effectively improve cognitive, social, physical, and affective outcomes. Despite these strengths, challenges in implementation and model fidelity, along with gaps in research on students with special needs, gender differences, and long-term effects, highlight the need for targeted interventions and further investigation (49).

This subtheme highlights the critical role of PE teacher professional development in enhancing student motivation and engagement in physical activity. While studies have consistently shown the positive impact of effective teaching methods, there remains a need for targeted interventions to address the challenges and limitations in implementation. The findings suggest that professional development opportunities, including workshops and lesson studies, can improve teachers’ autonomy support and instructional structure. However, further research is required to explore the long-term effects of these interventions and their effectiveness in addressing specific needs, such as gender differences and students with special needs. By investing in PE teacher professional development, we can create more engaging and motivating learning environments that foster students’ lifelong commitment to physical activity.

##### c. Family Support System

Research highlights the significant impact of family support on adolescents’ participation in physical activity (PA). For instance, strong family or parental support increases PA levels among girls, though logistic support from mothers is particularly effective (50, 51). Despite the clear role of family support internationally, evidence is scarce in Pakistan. A study in Poland found that while parental attitudes influence PA goals, they do not significantly affect children’s actual PA levels (52). In Australia, family support plays a crucial role in action sports, with parents and siblings providing essential encouragement and resources, particularly for girls in sports like mountain biking, skateboarding, and surfing (53). Tailored interventions are needed to address gender-specific support dynamics and enhance participation.

Studies underscore the significant role of family support in adolescents’ physical activity (PA). For example, research on dark triad personality traits in adolescent girls shows that psychopathy and Machiavellianism impact PA motivation both directly and indirectly, while narcissism affects motivation directly through increased self-focus (54). In Japan, direct logistical support from parents, such as enrolling children in sports clubs, was linked to higher levels of moderate-to-vigorous PA, whereas indirect support methods like modeling PA had less impact (55). A systematic review of family PA interventions found mixed results on overall family functioning but highlighted improvements in family cohesion and organization, especially in families with younger children (56). These findings emphasize the importance of targeted and logistical family support to enhance PA participation.

This subtheme highlights the crucial role of family support in promoting adolescents’ participation in physical activity. While research has consistently demonstrated the positive impact of family support in various contexts, the evidence in Pakistan remains limited. Studies from other countries have shown that family support can influence PA motivation, behavior, and overall well-being. However, the specific dynamics of family support and their effectiveness in different cultural contexts may vary. Further research is needed to explore the role of family support in promoting adolescent physical activity in Pakistan and to develop targeted interventions that address the unique needs and challenges of Pakistani families.

##### d. Role of Gender

Gender differences significantly impact physical activity (PA) among adolescents. In Brazil, boys predominantly engaged in sports like soccer and cycling, while girls participated in lower-intensity and less structured activities (57). Cultural factors further influence PA patterns. In Spain, cultural beliefs and discourses notably affect adolescent girls’ PA and their identity related to sports and physical education. Globally, boys are generally more active than girls and are more likely to meet recommended PA levels, as highlighted by the Global School Health Survey and a comparative study in low- and middle-income countries (58). A study across 49 low- and middle-income countries (LMICs) confirmed that low PA is more prevalent among adolescent girls (59).

Research from LMICs, including Pakistan, reveals how pro-male cultures and socio-cultural barriers limit girls’ PA, with public exercise often deemed inappropriate for females (36, 60). In Pakistan, gender significantly impacts physical activity (PA) levels. A discussion paper highlights that Pakistani girls often engage in PA at home and prefer lighter activities compared to boys, who participate in more strenuous sports like cricket and hockey (61).

Gender socialization in Pakistan reflects a patriarchal society, with boys engaging in fewer household chores and more active leisure activities, while girls are more likely to watch TV and face restricted access to public spaces (62). This issue is compounded by inadequate space for girls’ mobility in urban areas (63).

A study in Karachi found that girls aged 10-14 reported lower PA levels than boys, with a notable decline as they advanced in school grades. Cultural, religious, and contextual factors influenced PA, highlighting the need for targeted interventions to promote physical activity among Pakistani adolescents, especially girls (64). A study in Pakistan investigated barriers to women’s sports participation in urban and rural areas, finding economic and personal obstacles as the most significant barriers, with family and cultural factors being less impactful. The study suggests targeted advertising and diverse sports programs could improve participation and quality of life (65).

Another study in Lahore identified family influence, socio-cultural support, economic barriers, personal interest, and sexual harassment as key factors affecting women’s sports involvement, with economic barriers and sexual harassment having a notably negative impact. The study calls for collaborative efforts from policymakers, sports bodies, and communities to address these issues and increase female participation (66). In the context of social class, an analysis of high school soccer matches revealed that schools with higher proportions of working-class youth lost by larger margins, with less pronounced effects in predominantly Latinx schools. This indicates significant disparities linked to social class and community cultural wealth (67).

A qualitative review of barriers and facilitators to physical activity among young adult women highlighted time constraints, body image concerns, and family duties as barriers, while social support and community facilities were identified as facilitators. The review stresses the need for multilevel, culturally sensitive strategies (68). In Spain, research on university students found that fewer than half of both men and women were physically active, with women exhibiting lower levels of physical activity and higher external motivation compared to men. This underscores the need for interventions to address motivational differences (69). A study in the Netherlands explored how gender affects the use and perception of public open spaces for physical activity among adolescents. Boys and girls used these spaces differently, with girls favoring larger, well-maintained areas, suggesting the need for gender-sensitive design and maintenance of public spaces (70).

This subtheme highlights the significant impact of gender on adolescent physical activity levels, with boys generally more active than girls. Cultural factors, societal norms, and socioeconomic disparities play a crucial role in shaping these gender differences. In many contexts, girls face barriers such as limited access to public spaces, cultural restrictions, and economic constraints that hinder their participation in physical activity. Addressing these gender-specific challenges requires a multi-faceted approach that involves promoting gender equality, providing safe and inclusive spaces, and implementing targeted interventions to encourage girls’ engagement in physical activity.

##### e. Culturally Appropriate Dress

Culturally appropriate dress significantly affects physical activity (PA) participation, particularly for girls. A study noted that in Pakistani public schools, the absence of separate physical education (PE) uniforms forces students to wear regular clothing, leading to issues like skin infections due to heat and sweat (33). Similarly, another study highlighted that culturally and religiously appropriate dress is crucial for engaging girls in PA, suggesting separate uniforms and venues for boys and girls to enhance participation (60). In Indonesia, a study found that patriarchal views and objectification based on Islamic attire, like the veil, discourage Muslim female students from participating in PE and art activities. The study recommended separate PE classes as a temporary measure to boost participation (71).

In Pakistan, challenges related to dress codes, influenced by cultural and religious norms, hinder Muslim female athletes’ involvement in sports. Despite Islam’s endorsement of fitness, misconceptions and dress code restrictions limit participation, particularly in mixed-gender settings. Recommendations include addressing these dress code issues to better support female athletes and enhance their access to sport (72). A study in Ghana explored barriers to women’s and girls’ participation in community sports using Cooky and Messner’s theory of ‘the unevenness of social change.’ Through document analysis and interviews with sports officials, it identified that entrenched cultural norms and structural obstacles limit female participation and leadership in sports. The research suggests that enforcing gender policies could gradually drive cultural shifts and improve gender equality, but significant limitations persist (73).

This subtheme highlights the significant impact of culturally appropriate dress on adolescent physical activity participation, particularly for girls. In many contexts, cultural and religious norms dictate specific dress codes that can hinder female involvement in sports and physical education. This can lead to issues such as discomfort, embarrassment, and limited access to facilities. Addressing these challenges requires a multifaceted approach that involves promoting cultural sensitivity, providing appropriate facilities and equipment, and implementing policies that support gender equality in sports and physical activity.

##### f. Safety and Security

Safety and security concerns significantly impact outdoor physical activity (PA), particularly in developing countries. In Bangladesh, inadequate street lighting (62%) and lack of suitable PA spaces (56%) are major barriers for girls (74, 75). Similar issues are reported in India and South Africa, where safety concerns restrict girls’ PA (76). In Pakistan, safety fears lead parents to be overprotective, affecting girls’ engagement in PA (77, 78). In the US, a study of 1,437 adolescents found that access to school recreational facilities and public parks is linked to higher levels of moderate to vigorous physical activity (MVPA), with school facilities contributing an additional 4.4 minutes of daily MVPA on average (79).

This subtheme highlights the significant impact of safety and security concerns on adolescent physical activity, particularly in developing countries. Inadequate infrastructure, such as poor street lighting and lack of suitable PA spaces, can create barriers for girls and limit their opportunities for physical activity. Additionally, parental safety concerns can lead to overprotectiveness, further restricting girls’ engagement in outdoor activities. Addressing these safety concerns requires investment in infrastructure, promoting safe and inclusive spaces, and raising awareness about the importance of physical activity for adolescent health and well-being.

##### g. The Role of Infrastructure

Infrastructure significantly impacts physical activity (PA). A systematic review across 31 countries found that facilities like toilets, drinking fountains, and benches in public open spaces enhance their usability for PA (80). Organized sports clubs are positively linked to increased PA levels, with membership providing an average of four hours of exercise per week for many adolescents in Germany and a 60-69% increase in PA levels across six European countries (81, 82). In Pakistan, constraints such as limited resources and inadequate playground facilities hinder PA, particularly in private schools (8, 19, 63). A study found that walking facilities in neighborhoods improved PA levels (83). Additionally, a study in England identified “physical activity insecurity,” where marginalized youths experienced exclusion and discomfort in gyms and sports clubs, preferring natural spaces and youth clubs for their safety and comfort (84).

This subtheme highlights the significant impact of infrastructure on adolescent physical activity levels. Access to facilities such as toilets, drinking fountains, and benches in public open spaces can enhance their usability for PA. Organized sports clubs and well-maintained playgrounds can also provide opportunities for physical activity. However, in many developing countries, infrastructure constraints, such as limited resources and inadequate facilities, can hinder PA participation. Addressing these infrastructure challenges is essential to promoting physical activity among adolescents and creating inclusive and accessible spaces for everyone. This review primarily included sources available in English. As a result, this review may not include a few potentially relevant publications reflecting a wider global context in various other languages. The review was also limited to young adolescents from key databases. Some potentially pertinent publications may not have been identified with these search strategies or databases.

## Discussion

The primary research question for this review serves as a guide to this discussion with a focus on young adolescents’ physical activity levels and their influencing factors from a wider context including Pakistan. Research on physical activity levels among young adolescents has been carried out extensively, and recommended levels of physical activity are not met in high, middle, and low-income countries alike. However, little is known about the factors that might influence physical activity levels among young adolescent students in low-resourced countries such as Pakistan. PA believed it to be less important than academic studies. Priority is thus given to academic studies (6, 7). Overall, there is evidence to indicate that young adolescents’ physical activity levels declined with age due to several socio-cultural and environmental factors. Very little attention has been paid to understanding the factors influencing the physical activity of secondary school adolescents in a cultural context. This review’s major research topic guides this debate, which focuses on the physical activity levels of adolescents and their affecting elements from a broader perspective, including Pakistan. There is evidence that the physical activity levels of adolescents decrease with age owing to many sociocultural and environmental influences (85). Understanding the cultural elements that influence the physical activity of secondary school teenagers has received very little study. This research indicated a lack of physical education policies and guidelines as one of the causes of adolescents’ lack of physical exercise. Unfortunately, there are nations, such as Pakistan, where these norms do not exist. In contrast, nations with physical activity rules and standards continue to fall short of the recommended levels of physical activity.

There are complications in implementing these rules. Secondly, there seems to be evidence of differences in the general physical education recommendations for secondary school students in the literature.

Therefore, the generalization of such published literature on implementing physical education guidelines can be problematic because these guidelines were proposed without taking into account key factors, such as age and gender, that influence physical activity among boys and girls in various settings. In addition, since boys and girls have distinct developmental trajectories, the usual criteria for physical education in terms of hours allotted in a curriculum may not work for both sexes in various circumstances (11). The importance of performing research to address the identified information gaps on physical activity among early teenage pupils is outlined by these studies. It stressed the need to adopt school-based physical education guidelines and policies to further this initiative to increase physical activity among this generation of teenagers.

The separate uniform for PA was identified as a major concern, and therefore, it was suggested that separate uniforms and venues should be considered for boys and girls to increase engagement in PA, a review of literature by Abbasi also pointed out that sometimes wearing a culturally appropriate dress has a special connotation from a religious perspective. For example, in Muslim culture, dressing in a manner that reveals body parts may not be considered culturally appropriate (60).

A systematic review and meta-analysis were conducted from March 2019 to April 2019 to determine the prevalence of physical activity among adolescent students in Pakistan. The results showed high heterogeneity in reporting the proportion of physically active adolescents from 6% to 70%. Therefore, this review has presented the pooled prevalence of adolescents’ physical activity as 36%, almost twice the prevalence of physical activity estimated by the Pakistan Global School Health Survey (15.5%) (16). This review included the studies irrespective of low-quality indicators. Besides this, the studies included have not adjusted key confounding variables, and therefore, generalizability could not be guaranteed.

Further, both boys and girls have different developmental trajectories; therefore, the standard guidelines for physical education in terms of hours allocation in a curriculum may not work for both sexes in different contexts. These studies collectively outline a critical role for conducting a study in addressing the knowledge gaps on physical activity among young adolescent students identified through this review. It requires further research to understand it. The review was also limited to young adolescents from key databases. Some potentially pertinent publications may not have been identified with these search strategies or databases.

### Recommendations for Future Practice

This study highlights the need for policy development to integrate physical education (PE) into school settings in Pakistan. Close coordination between the health and education ministries is essential to ensure widespread access to physical activity (PA) opportunities for adolescents. Establishing well-equipped gymnasiums and implementing safety measures are crucial for fostering a secure PA environment.

Schools should incorporate health education programs targeting students, parents, teachers, and policymakers. Professional development for PE teachers must be prioritized to enhance teaching skills and PA promotion. A designated PA uniform made of non-transparent, weather-appropriate material should be introduced.

To support girls’ participation, self-defense training should be offered, both within and outside of schools. Policies must ensure sustainable PA opportunities for girls, while societal awareness campaigns should promote gender equality in sports and PA.

### Recommendations for Future Research

This study focused on understanding adolescent PA. Future research should explore the perspectives of education and health ministry leaders, as well as school management, on implementing school-based preventive health programs. Engaging policymakers is crucial for driving national-level changes in PA promotion.

There is a significant gap in research on adolescent girls’ PA in Pakistan. Studies should examine ways to promote their participation through insights from parents and teachers, with a focus on gender dynamics. A cross-national study comparing public and private schools would provide deeper insights into PA trends. Further research on policy implementation and evaluation in school settings is also needed.

## Conclusion

This comprehensive scoping review highlights the multifaceted factors influencing adolescent physical activity levels in Pakistan. The findings reveal a consistent decline in physical activity as children transition into adolescence, with gender, cultural norms, and socioeconomic disparities playing significant roles. While physical activity offers numerous benefits for adolescent health and well-being, various barriers, including limited resources, infrastructure constraints, and societal norms, hinder their engagement.

To promote physical activity among Pakistani adolescents, it is essential to address these challenges through targeted interventions. This includes enhancing physical education curricula, providing professional development for PE teachers, fostering family support, promoting gender equality, and investing in infrastructure development. By implementing these strategies, we can create a more conducive environment for adolescents to engage in regular physical activity and reap the associated benefits.

## Data Availability

N/A

## Declarations

### Ethical consideration & consent to participate

As this study is a scoping review, it did not involve direct human or animal participants and, therefore, did not require formal ethical approval or informed consent. However, all included studies were reviewed ethically and adhered to the principles of research integrity. This review was conducted in accordance with the guidelines outlined in the Declaration of Helsinki, ensuring that the ethical considerations of the primary studies included in the review were respected.

### Consent for Publication

Not applicable

### Availability of data and materials

As this study is a scoping review, no primary data were generated or collected. All data analyzed in this review were obtained from publicly available sources, including peer-reviewed journal articles and grey literature. The full list of included studies and relevant materials are available from the corresponding author upon reasonable request.

### Authors Contribution

**SG** conceptualized the study, designed the research framework, developed the scoping review protocol, conducted the literature search, screened and selected studies, performed data extraction and synthesis, and contributed to manuscript writing and revisions. **JMA** assisted in study design, contributed to data extraction and synthesis, provided methodological guidance for the scoping review process, critically reviewed the manuscript for intellectual content, and contributed to finalizing the manuscript. **SA** contributed to the literature search, screening, and data extraction, assisted in organizing the results, and provided input on the discussion section. **AA** supported data extraction and synthesis, assisted in manuscript writing and editing, and contributed to refining the final version of the manuscript.

## Acknowledgement

This PhD thesis, authored by **SG**, is dedicated to expressing our deep appreciation to all co-authors for their invaluable guidance and support throughout this research.

We would like to express our gratitude to Aga Khan University for providing the necessary resources and support for conducting this scoping review. We also acknowledge the invaluable assistance of the university librarians in literature research, data organization, and technical support.

Additionally, we extend our sincere appreciation to our colleagues and peers for their constructive feedback, which significantly contributed to improving this manuscript.

